# Assessment of Bedside-Adaptable Models to Predict Molecular Sepsis Subtypes in a Resource-Limited Setting: A Multicenter Analysis from Uganda

**DOI:** 10.64898/2026.04.08.26350396

**Authors:** Barnabas Bakamutumaho, Julius J. Lutwama, Nicholas Owor, Xuan Lu, Peter James Eliku, Joyce Namulondo, John Kayiwa, Jesse E. Ross, Christopher Nsereko, John Bosco Nsubuga, Joseph Shinyale, Ignatius Asasira, Tonny Kiyingi, Steven J. Reynolds, Kai Nie, Seunghee Kim-Schulze, Matthew J. Cummings

## Abstract

**Objective:** Biologically defined sepsis subtypes have been identified in low- and middle-income countries (LMICs), but limited access to molecular diagnostics challenges broader evaluation and implementation in resource-limited settings. We assessed whether models including bedside clinical and rapid microbiologic data could accurately stratify Ugandan adults with sepsis by molecular subtype.

**Design:** Secondary analysis of two prospective observational sepsis cohorts, testing bedside-adaptable classifier models against transcriptomic and proteomic subtype assignments.

**Setting:** Entebbe Regional Referral Hospital (urban) and Tororo General Hospital (rural), Uganda.

**Patients:** Adults (≥18 years) hospitalized with sepsis, with available transcriptomic (N=355) and/or proteomic (N=495) profiling enabling subtype assignment.

**Interventions:** None.

**Measurements and Main Results:** Using data from two prospective cohorts (RESERVE-U-2-TOR and RESERVE-U-1-EBB), we evaluated bedside-adaptable models against Uganda-derived molecular sepsis subtypes, and, secondarily, against molecular subtypes and axes derived in high-income countries. In RESERVE-U-2-TOR, clinical models including demographics and bedside physiological variables demonstrated moderate discrimination for transcriptomic and proteomic subtype assignment (AUROC 0.75 [95% CI, 0.69–0.81] and 0.73 [0.66–0.80], respectively) with strong calibration (Integrated Calibration Index [E_avg_] ≤0.015 for both models). Adding rapid diagnostic results for HIV, malaria, and tuberculosis produced similar performance (AUROC 0.76 and 0.74; E_avg_ ≤0.016). In RESERVE-U-1-EBB, discrimination for clinical and clinico-microbiological models was more variable (AUROC range 0.63–0.75) while calibration remained acceptable (E_avg_ ≤0.053). Performance was similar when models were evaluated against molecular sepsis frameworks derived in high-income countries, with acceptable calibration and moderate discrimination.

**Conclusions:** Bedside-adaptable clinical models, with or without rapid microbiologic testing, demonstrated acceptable calibration but only modest discrimination for molecular sepsis subtype assignment in Uganda. Expanding laboratory capacity and access to scalable, low-cost molecular biomarker assays will be necessary to advance precision sepsis care in LMIC settings.

**Key Points:** *Question:* Among adults hospitalized with sepsis in a resource-limited setting, can bedside clinical variables, alone or combined with rapid pathogen diagnostics, accurately stratify molecular sepsis subtype assignments?

*Findings:* In two prospective Ugandan sepsis cohorts, bedside clinical and clinico-microbiologic models showed robust calibration but only modest discrimination for classifying Uganda-derived transcriptomic and proteomic subtypes. Models also achieved moderate performance for stratifying high-income-country-derived transcriptomic subtypes and immune dysfunction axes, suggesting bedside variables reflect illness severity but incompletely capture underlying molecular signatures.

*Meaning:* Bedside-adaptable models can support reasonably calibrated risk estimation for molecular sepsis stratification in resource-limited settings but lack sufficient discriminatory power to serve as stand-alone tools. These findings support efforts to improve acute-care laboratory capacity and access to scalable molecular biomarker panels, with the goal of enabling precision sepsis care in low- and middle-income countries.

## Introduction

Over the past decade, molecular host-response profiling in high-income countries (HIC) has established biological subtypes of sepsis and related critical illness that are associated with distinct clinical trajectories, outcomes, and treatment responses (1-6). To support clinical application, models incorporating routinely available clinical and laboratory data have been developed to assign patients to molecular subtypes, supporting the feasibility of rapid, near-bedside assignment in HIC settings (7,8).

In sub-Saharan Africa, where the global sepsis burden is concentrated, we recently derived and validated molecular sepsis subtypes using transcriptomic and proteomic profiling of Ugandan sepsis cohorts (9-11). Across both omics platforms, we identified two reproducible subtypes distinguished by differential activation of inflammatory and endothelial pathways and associated with differences in physiological severity and survival (10,11). Despite these advances, translation in sub-Saharan Africa and other low- and middle-income country (LMIC) settings is constrained by limited access to omics platforms and molecular biomarker assays. We therefore sought to determine whether bedside-adaptable models could stratify Ugandan adults with sepsis by molecular subtype, enabling biologically informed triage and risk stratification in settings where advanced molecular testing is not feasible.

## Methods

### Study Setting and Participants

We analyzed data from two prospective cohorts of adults hospitalized with sepsis in Uganda: RESERVE-U-2-TOR (Tororo District Hospital [rural], 2021–2023) and RESERVE-U-1-EBB (Entebbe Regional Referral Hospital [urban], 2017–2019). Enrollment criteria were uniform across RESERVE-U cohorts, details of which have been published (10,11). Briefly, non-pregnant adults (≥18 years) were eligible if they had suspected infection (history of fever or axillary temperature ≥37.5°C) and illness severe enough to warrant hospital admission. For omics profiling, we included patients at higher risk of poor outcomes typical of sepsis, defined as those with a quick Sepsis-related Organ Failure Assessment (qSOFA) score ≥1 (respiratory rate ≥22/min, systolic blood pressure ≤100 mm Hg, or altered mentation) (12). As others in the region have done, we chose this cutoff because it is consistently predictive of mortality in LMICs with greater sensitivity than qSOFA score ≥2 (13-15).

### Outcome Definitions

For the analyses described herein, the outcome in each model was assignment to molecular subtypes previously derived and validated through omics analyses of the RESERVE-U cohorts (10,11). Uganda-derived transcriptomic subtypes (Uganda Sepsis Endotypes [USE]-1/USE-2) were identified by unsupervised clustering of whole-blood RNAseq data, and Uganda-derived proteomic subtypes (Uganda Sepsis Signature [USS]-1/USS-2) were derived using similar methods applied to Olink serum proteomic data (10,11).

In secondary analyses, we evaluated bedside-adaptable models against consensus transcriptomic subtypes and continuous host-response axes recently derived and validated in HICs, including Consensus Transcriptomic Subtypes (CTS) and Hi-DEF myeloid and lymphoid dysregulation scores (4,5). CTS assignments and Hi-DEF scores were generated using published algorithms (4,5).

### Predictor Selection and Modeling

Among clinical and microbiologic variables prospectively collected in the RESERVE-U cohorts, predictors were chosen *a priori* based on bedside availability, differential associations with subtype assignments, and clinical and biological relevance, and were used as inputs for two nested models to predict molecular subtypes. The clinical model included age, sex, duration of illness prior to study enrollment (i.e., sample collection), temperature, heart rate, respiratory rate, systolic blood pressure, oxygen saturation, and mental status, the latter as per the “Alert, Verbal, Pain, and Unresponsive” consciousness scale. The clinico-microbiologic model additionally included results of rapid diagnostic testing for HIV, malaria, and tuberculosis (TB; urine TB-LAM and/or sputum Xpert MTB/RIF Ultra), performed in the RESERVE-U cohorts as previously described (10,11). Testing for these pathogens was informed by regional epidemiology and World Health Organization guidelines for management of sepsis and septic shock in sub-Saharan Africa (16,17).

We evaluated the extent to which routinely available bedside variables capture molecular subtype structure across three outcome types: (1) binary Uganda-derived subtypes (USE-1/USE-2; USS-1/USS-2), (2) multiclass categorical subtypes (CTS1/CTS2/CTS3), and (3) continuous host-response axes (Hi-DEF myeloid and lymphoid dysregulation scores). All analyses were performed in R (v4.3) via RStudio using base R functions (*stats*) and additional packages as described below. For binary outcomes, we fit multivariable logistic regression models (*stats*). For CTS, we fit multinomial logistic regression models treating CTS as an unordered three-class outcome (*nnet*). For continuous Hi-DEF scores, we fit multivariable linear regression models (*stats*). In the linear regression analyses, coefficient inference was supplemented using heteroskedasticity-robust variance estimators (*sandwich*) with testing via *lmtest*. Across modeling approaches, multicollinearity was assessed using variance inflation factors (VIF; *car*), and overall collinearity was further evaluated using the condition number derived from the predictor matrix.

Our primary objective was to assess, within each cohort, how accurately bedside variables predicted molecular subtypes or axes, rather than to develop a single transportable prediction tool. Accordingly, we fit models independently within each cohort using the same pre-specified covariates. For binary outcomes, discrimination was assessed using the area under the receiver-operating-characteristic curve (AUROC; *pROC*), with 95% confidence intervals (CI) obtained by bootstrap resampling (10,000 iterations). Calibration was assessed using LOESS-based calibration models and summarized by the Integrated Calibration Index (E_avg_), defined as the average absolute difference between predicted probabilities and LOESS-smoothed observed probabilities, with lower values indicating better calibration (18). For CTS, performance was assessed using multiclass AUROC (*pROC*) with 95% CIs obtained by bootstrap resampling (10,000 iterations), together with macro-averaged LOESS-based Integrated Calibration Index. For continuous Hi-DEF outcomes, linear regression model performance was summarized using the Pearson correlation between observed and predicted myeloid and lymphoid dysregulation scores, with 95% CIs obtained by bootstrap resampling (10,000 iterations), as well as mean absolute error (MAE).

### Ethics approval and reporting statement

In both RESERVE-U cohorts, each enrolled participant or their surrogate provided written informed consent. The study protocol (“Clinical and molecular epidemiology of severe acute febrile and respiratory illness in Uganda”) was approved by ethics committees at Columbia University (AAAR1450 on December 13^th^, 2016) and Uganda Virus Research Institute (GC/127/17/02/582 on February 3^rd^, 2017). All study procedures were followed in accordance with the ethical standards of the responsible institutional committees and the Helsinki Declaration of 1975. Results are reported in accordance with the Transparent Reporting of a multivariable prediction model for Individual Prognosis or Diagnosis (TRIPOD) checklist for prediction models (19).

## Results

### Cohort Characteristics

Across both cohorts, we analyzed data from 495 participants assigned to proteomic subtypes (RESERVE-U-2-TOR N=253; RESERVE-U-1-EBB N=242) and 355 participants assigned to transcriptomic subtypes (RESERVE-U-2-TOR N=243; RESERVE-U-1-EBB N=112) (**Tables S1-S4**). Reflective of the epidemiology of sepsis in sub-Saharan Africa, participants were primarily younger adults with a high prevalence of HIV co-infection. Severe malaria and TB were common, and mortality was substantial (27-30%).

As previously described, we derived and validated two molecular subtypes through transcriptomic (USE) and proteomic (USS) analyses of the RESERVE-U cohorts (10,11) (**Tables S1-S4**). The higher-risk subtypes (USE-2 and USS-2) were characterized by pro-inflammatory innate immune activation, lymphocyte suppression, and microvascular dysfunction, and were clinically associated with increased physiological severity and higher mortality. USE-2 and USS-2 were also associated with higher frequencies of HIV and TB, whereas USE-1 and USS-1 were more frequently associated with malaria.

Across both cohorts, CTS demonstrated variable clinical associations. In RESERVE-U-2-TOR, both CTS1 and CTS2 were associated with greater physiological severity and higher mortality than CTS3. In RESERVE-U-1-EBB, CTS1 was associated with greater illness severity and higher mortality, whereas CTS2 and CTS3 showed comparably lower severity and mortality (**Tables S5-S6**). Hi-DEF axes demonstrated graded, albeit variable, relationships with clinical severity (**Tables S7-S10**). In RESERVE-U-2-TOR, higher lymphoid dysregulation scores were associated with greater physiological severity and higher mortality, and increasing myeloid dysregulation scores tracked with greater clinical severity. In RESERVE-U-1-EBB, higher lymphoid dysregulation scores were again associated with greater severity and higher mortality, whereas myeloid dysregulation scores showed weaker associations.

### Classifier performance for stratification of Uganda-derived molecular subtypes

Multicollinearity among predictors was assessed prior to model interpretation using VIFs and condition indices derived from the predictor matrix; no evidence of problematic collinearity was identified (all VIFs < 1.75). Across RESERVE-U-2-TOR, both the clinical and clinico-microbiological models showed moderate discrimination for transcriptomic subtypes (USE), with AUROCs of 0.75 and 0.76, respectively (**Figure 1, Table S11 in Supplement**). Calibration was strong for both models (E_avg_ ≤0.016). For proteomic subtypes (USS) in RESERVE-U-2-TOR, discrimination was similar (AUROC 0.73 for clinical vs 0.74 clinico-microbiological models), as was calibration (**Figure 2, Table S11 in Supplement**).

**Figure 1.**
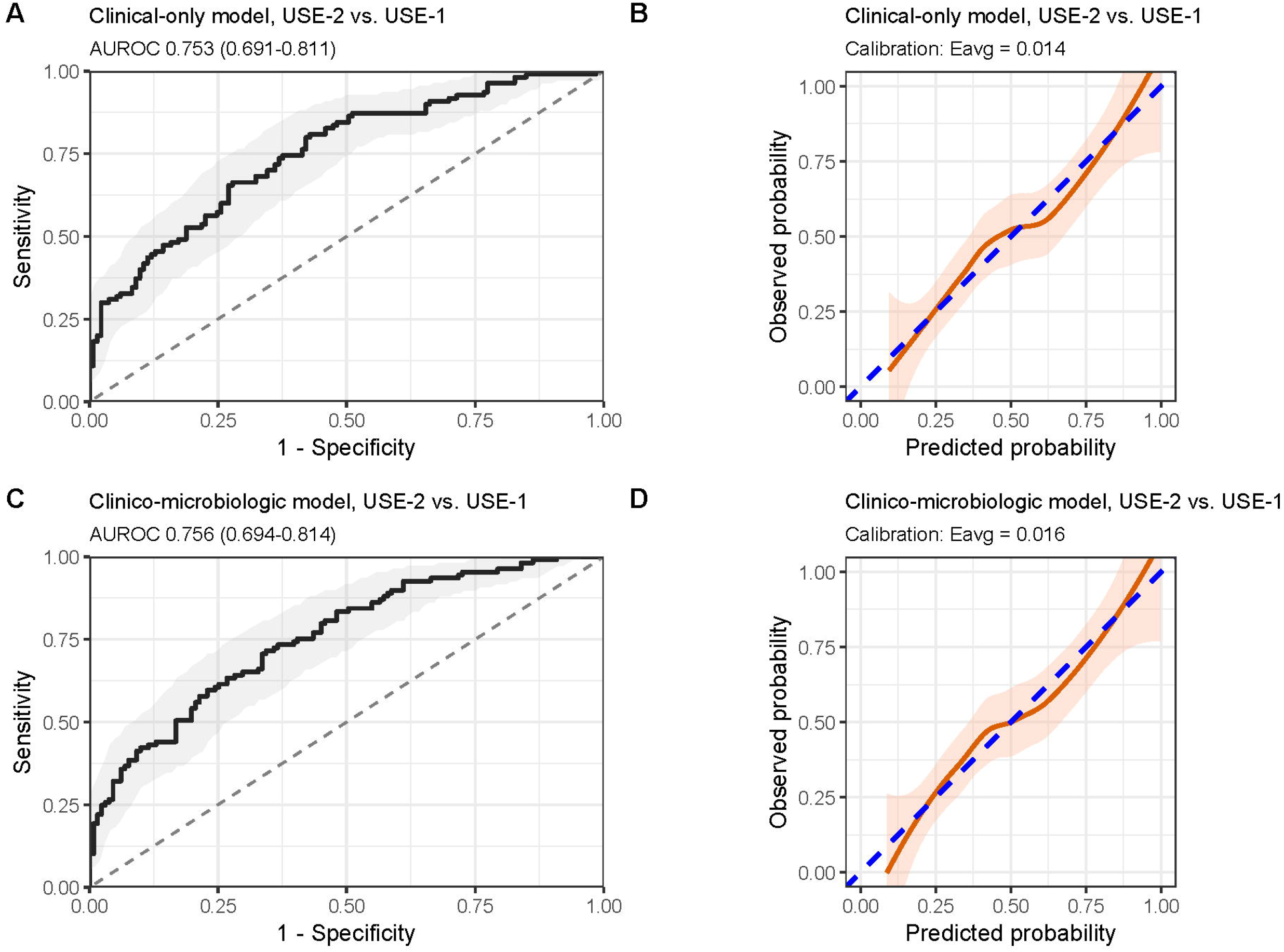
Discrimination and calibration of clinical-only (N=243) and clinico-microbiologic (N=240) models for prediction of Uganda-derived transcriptomic subtypes (Uganda Sepsis Endotypes [USE]) in the RESERVE-U-2-TOR cohort. **(A)** Receiver operating characteristic (ROC) curve for the clinical-only model distinguishing USE-2 from USE-1, with shaded 95% confidence interval (CI) band. **(B)** Calibration plot for the clinical-only model showing loess-smoothed observed versus predicted probability; the dashed diagonal indicates perfect calibration. **(C)** ROC curve for the clinico-microbiologic model distinguishing USE-2 from USE-1, with shaded 95% CI band. **(D)** Calibration plot for the clinico-microbiologic model showing loess-smoothed observed versus predicted probability; the dashed diagonal indicates perfect calibration. AUROC values with 95% CI bands are shown on the ROC panels, and calibration error is summarized by the Integrated Calibration Index (i.e., estimated average calibration error [Eavg]) on the calibration panels.

**Figure 2.**
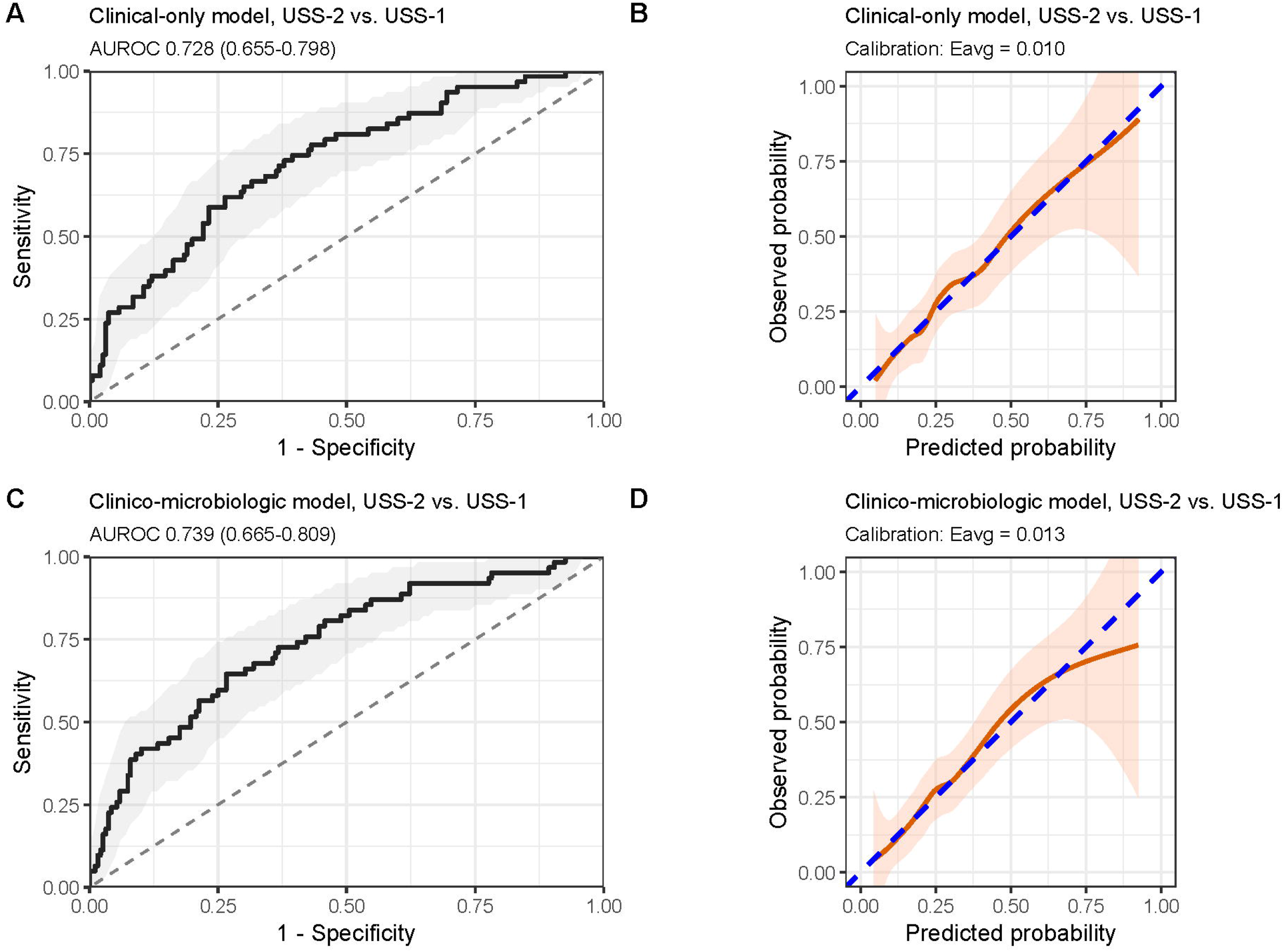
Discrimination and calibration of clinical-only (N=253) and clinico-microbiologic (N=250) models for prediction of Uganda-derived proteomic subtypes (Uganda Sepsis Signatures [USS]) in the RESERVE-U-2-TOR cohort. **(A)** Receiver operating characteristic (ROC) curve for the clinical-only model distinguishing USS-2 from USS-1, with shaded 95% confidence interval (CI) band. **(B)** Calibration plot for the clinical-only model showing loess-smoothed observed versus predicted probability; the dashed diagonal indicates perfect calibration. **(C)** ROC curve for the clinico-microbiologic model distinguishing USS-2 from USS-1, with shaded 95% CI band. **(D)** Calibration plot for the clinico-microbiologic model showing loess-smoothed observed versus predicted probability; the dashed diagonal indicates perfect calibration. AUROC values with 95% CI bands are shown on the ROC panels, and calibration error is summarized by the Integrated Calibration Index (i.e., estimated average calibration error [Eavg]) on the calibration panels.

In RESERVE-U-1-EBB, model performance was more heterogeneous. For USE assignments, discrimination declined relative to RESERVE-U-2-TOR (AUROC 0.63 for clinical model) but improved with microbiologic variables (AUROC 0.69) (**Figure 3, Table S12 in Supplement**). Calibration remained reasonable (E_avg_ ≤0.053 for both models). Performance for USS classification was relatively preserved, with AUROCs of 0.72 and 0.75 for clinical and clinico-microbiological models, respectively, alongside improved calibration, together indicating better discrimination and more reliable risk estimates when microbiological variables were included (**Figure 4, Table S12 in Supplement**).

**Figure 3.**
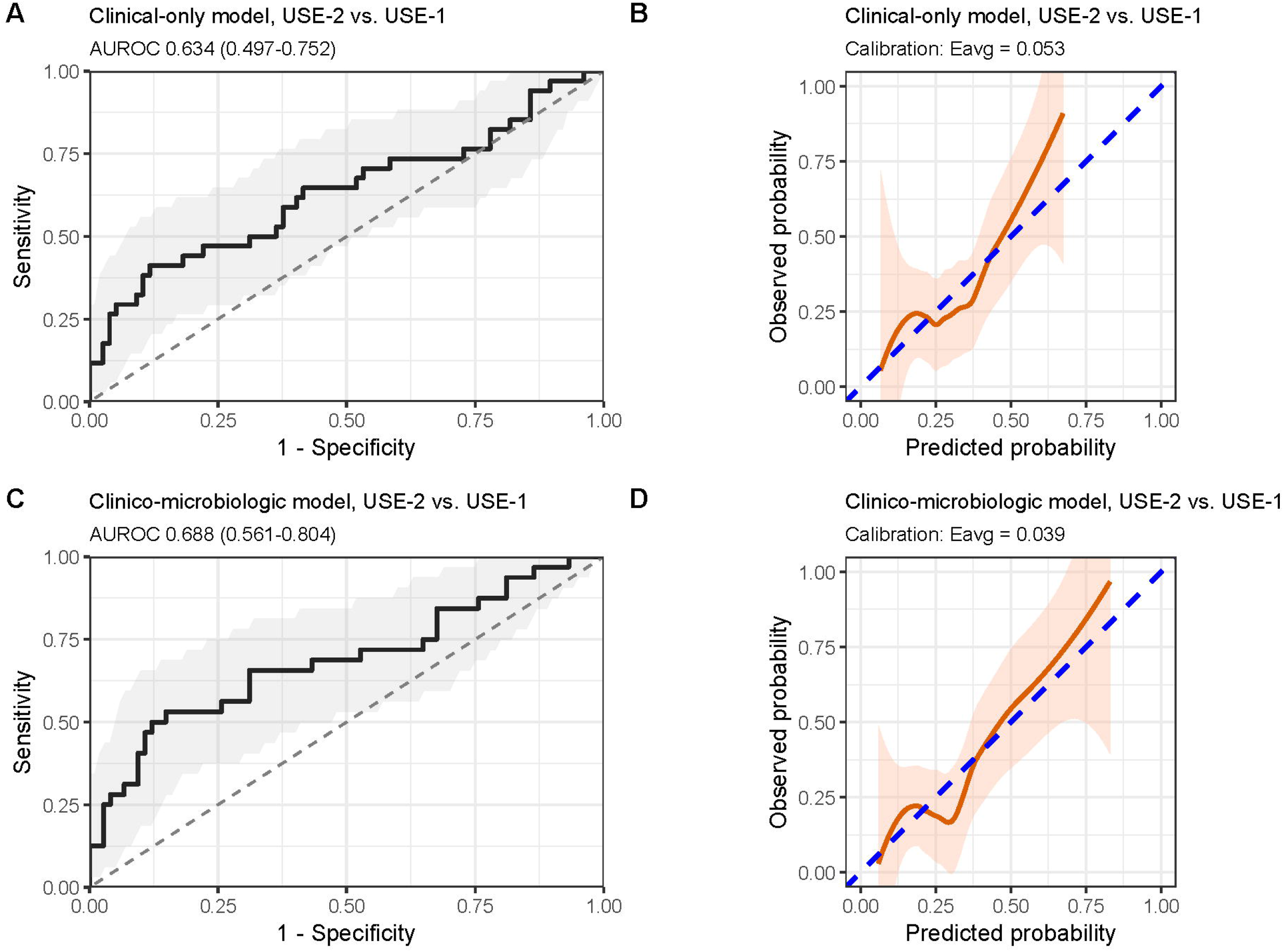
Discrimination and calibration of clinical-only (N=111) and clinico-microbiologic (N=106) models for prediction of Uganda-derived transcriptomic subtypes (Uganda Sepsis Endotypes [USE]) in the RESERVE-U-1-EBB cohort. **(A)** Receiver operating characteristic (ROC) curve for the clinical-only model distinguishing USE-2 from USE-1, with shaded 95% confidence interval (CI) band. **(B)** Calibration plot for the clinical-only model showing loess-smoothed observed versus predicted probability; the dashed diagonal indicates perfect calibration. **(C)** ROC curve for the clinico-microbiologic model distinguishing USE-2 from USE-1, with shaded 95% CI band. **(D)** Calibration plot for the clinico-microbiologic model showing loess-smoothed observed versus predicted probability; the dashed diagonal indicates perfect calibration. AUROC values with 95% CI bands are shown on the ROC panels, and calibration error is summarized by the Integrated Calibration Index (i.e., estimated average calibration error [Eavg]) on the calibration panels.

**Figure 4.**
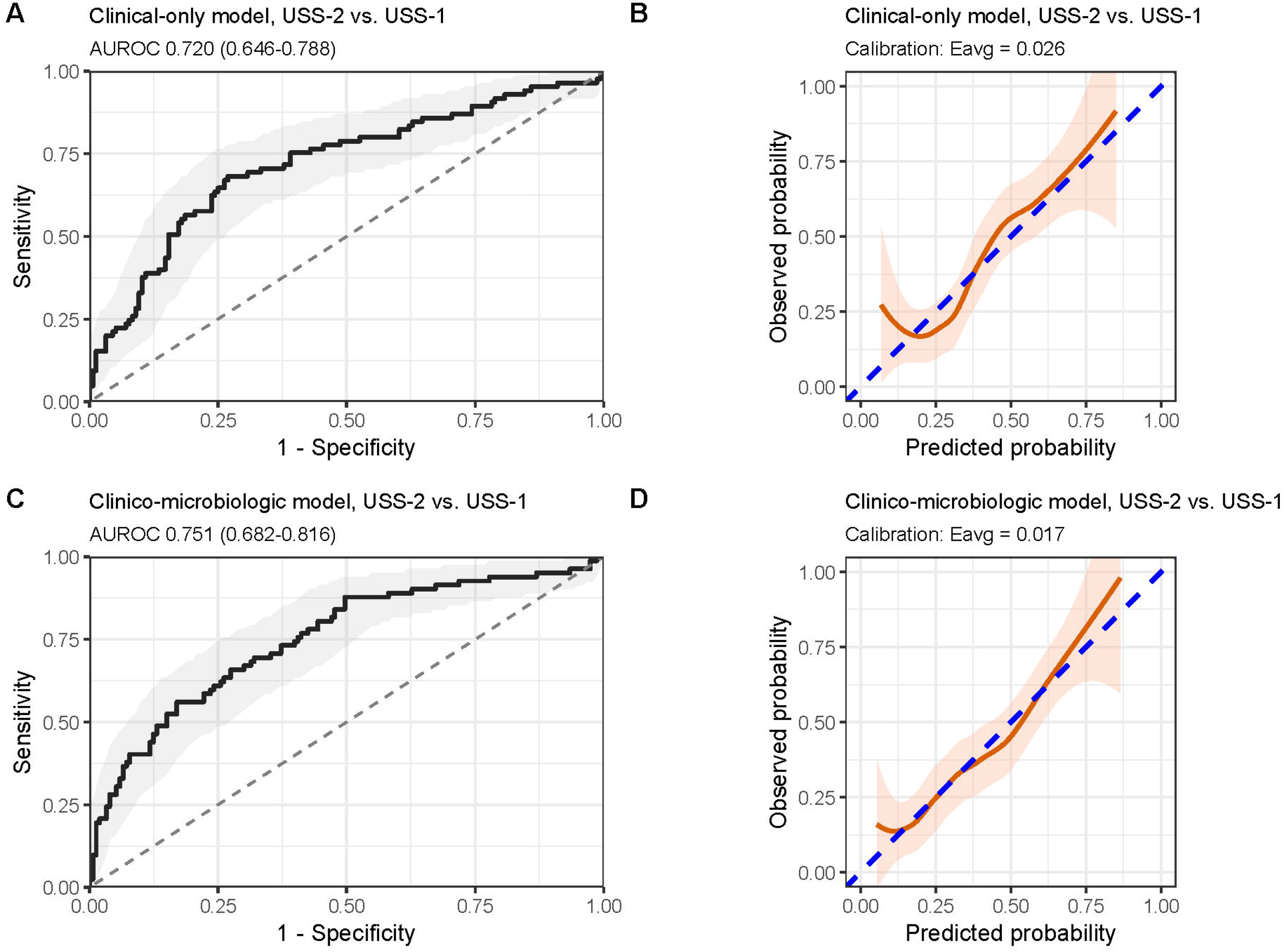
Discrimination and calibration of clinical-only (N=241) and clinico-microbiologic (N=235) models for prediction of Uganda-derived proteomic subtypes (Uganda Sepsis Signatures [USS]) in the RESERVE-U-1-EBB cohort. **(A)** Receiver operating characteristic (ROC) curve for the clinical-only model distinguishing USS-2 from USS-1, with shaded 95% confidence interval (CI) band. **(B)** Calibration plot for the clinical-only model showing loess-smoothed observed versus predicted probability; the dashed diagonal indicates perfect calibration. **(C)** ROC curve for the clinico-microbiologic model distinguishing USS-2 from USS-1, with shaded 95% CI band. **(D)** Calibration plot for the clinico-microbiologic model showing loess-smoothed observed versus predicted probability; the dashed diagonal indicates perfect calibration. AUROC values with 95% CI bands are shown on the ROC panels, and calibration error is summarized by the Integrated Calibration Index (i.e., estimated average calibration error [Eavg]) on the calibration panels.

### Classifier performance for stratification of HIC-derived molecular subtypes and axes

Performance of clinical and clinico-microbiological models was also modest for HIC-derived transcriptomic frameworks. For CTS assignments in RESERVE-U-2-TOR, discrimination and calibration for clinical and clinico-microbiological models were similar (AUROC 0.68 and E_avg_ 0.018-0.019) (**Table S11 in Supplement**). In RESERVE-U-1-EBB, adding microbiologic variables more clearly improved discrimination for CTS assignments (AUROC 0.69 to 0.74) while calibration was unchanged (**Table S12 in Supplement**). For Hi-DEF myeloid dysregulation scores in RESERVE-U-2-TOR and RESERVE-U-1-EBB, the clinical-only and clinico-microbiological models performed similarly and achieved weak positive correlations with observed scores (*r* = 0.28-0.29 and *r* = 0.26-0.28, respectively), with MAE values suggesting moderate prediction error relative to score distributions (**Tables S11-S12 in Supplement**). For lymphoid dysregulation scores, correlations were higher though remained moderate in RESERVE-U-2-TOR (*r* = 0.52 for both models) and RESERVE-U-1-EBB (*r =* 0.40 and 0.49 for clinical and clinico-microbiological models, respectively), along with lower MAE values relative to lymphoid score distributions.

## Discussion

Sepsis is associated with a disproportionate burden of morbidity and mortality in LMICs, yet the molecular tools most commonly used to advance precision sepsis care are unavailable in nearly all frontline settings (20,21). In HICs, multiple groups have shown that molecular subtypes can be classified using routinely collected clinical and laboratory data (including electronic medical record [EMR]-sourced variables), with generally strong performance (7,8). In this context, we evaluated whether bedside-adaptable models, restricted to data realistically available near the bedside in resource-limited settings, could stratify Ugandan adults with sepsis by molecular subtypes and host-response axes.

Overall, bedside clinical and clinico-microbiologic models demonstrated robust calibration but only modest discrimination for both Uganda-derived molecular subtypes and HIC-derived frameworks. These findings suggest that although physiological variables reflect downstream manifestations of illness severity, they do not fully capture the underlying pathobiological programs represented by transcriptomic and proteomic signatures. The addition of rapid pathogen diagnostics yielded incremental improvement for several molecular endpoints, but these gains were generally modest. This likely reflects the limited biological resolution of qualitative microbiologic measures; although essential for antimicrobial management, they may not reliably distinguish host-response programs underlying pathobiological heterogeneity. Together, these findings highlight an important implementation gap in extending precision sepsis care to LMIC settings; clinical-only models may be feasible and reasonably well calibrated, but they often lack sufficient discriminatory power to serve as stand-alone tools for molecular subtype assignments. Our results therefore underscore the need to expand access to foundational acute care laboratory testing, including basic chemistry and hematology. They also support development of parsimonious biomarker panels and integrated clinical, microbiologic, and molecular approaches that better capture sepsis pathobiology and can be deployed on scalable rapid platforms, such as targeted quantitative polymerase chain reaction and immunoassays (21).

Across cohorts, the molecular strata most consistently associated with worse outcomes were our Uganda-derived subtypes, alongside selected HIC-derived frameworks, most notably CTS1 and the Hi-DEF lymphoid dysregulation score. The prognostic relevance of CTS2 and the Hi-DEF myeloid dysregulation score appeared more variable, underscoring a practical trade-off between using broader consensus subtyping to enable cross-cohort comparability versus adopting regionally tailored molecular definitions that may better capture locally dominant pathobiology and clinical risk. These findings of prognostic enrichment may inform the design of future sepsis trials in sub-Saharan Africa (e.g., of supportive care and host-directed therapeutics), that assess heterogeneity of treatment effect by molecular strata, prioritizing subgroup analyses by reproducibly meaningful signatures (22-24).

This work has limitations. First, our models were intentionally constrained to predictor sets plausibly available near the bedside in resource-limited settings; this pragmatic design improves feasibility but likely caps discrimination for biologically complex molecular subtypes. Second, because our primary goal was to quantify how strongly bedside variables discriminate molecular subtypes/axes (rather than to develop a single transportable tool), models were fit separately within each cohort using the same pre-specified covariates, rather than deriving coefficients in one cohort and applying them unchanged to the other; accordingly, these results do not directly estimate out-of-cohort performance of a fixed model. Third, in RESERVE-U-1-EBB, transcriptomic data were available for a subset (approximately 40%) of patients; although the characteristics of this subset were similar to those of the full cohort, selection bias cannot be excluded, and the smaller sample size may have contributed to the lower observed model performance and less precise estimates (10). Fourth, cohort differences (case mix, pathogen distribution, comorbidities such as HIV, and timing of presentation) may affect pathological and clinical signals used for prediction, limiting transportability across sites and stressing the need for regional recalibration.

In Ugandan adults with sepsis, bedside-adaptable models showed strong calibration but only modest discrimination for Uganda-derived and HIC-derived molecular subtype frameworks. These findings underscore both the promise and current limitations of purely clinical stratification approaches in LMIC settings and support investment in scalable, rapid molecular and biomarker platforms, integrated with clinical and microbiologic data, to enable equitable, biologically informed precision sepsis care.

## Data Availability

: RNAseq data from the RESERVE-U-2-TOR cohort are available in the NIH/NCBI Sequence Read Archive through dbGaP under accession number phs003914.v1.p1. In concordance with participant consent and institutional certification of genomic data sharing, RNAseq data from RESERVE-U-2-TOR will be available to investigators with an IRB-approved protocol. Requests for access can be made to the NIH/NIAID Data Access Committee. RNAseq data from the RESERVE-U-1-EBB cohort are available in the NIH/NCBI Sequence Read Archive under accession number PRJNA794277. Proteomic data from RESERVE-U-2-TOR and RESERVE-U-1-EBB are available in Dryad at https://datadryad.org/dataset/doi:10.5061/dryad.b2rbnzsq2. Other de-identified data will be made available to researchers affiliated with an appropriate institution following mutual signing of a data access agreement and obtainment of necessary ethics approvals.

