## Supplemental Materials for "Assessment of Bedside-Adaptable Models to Predict Molecular Sepsis Subtypes in a Resource-Limited Setting: A Multicenter Analysis from Uganda"

**Supplemental Digital Content**

**Table S1: Patient characteristics stratified by Uganda Sepsis Endotypes (USE) in RESERVE-U-2-TOR**

| **Characteristic** | **N** | **Overall**  **N = 243*^1^*** | **USE-1**  **N = 133*^1^*** | **USE-2**  **N = 110*^1^*** | **p-value*^2^*** |
| --- | --- | --- | --- | --- | --- |
| Age, years | 243 | 48 (35, 62) | 45 (30, 58) | 50 (40, 70) | 0.004 |
| Female sex | 243 | 147/243 (60.5%) | 87/133 (65.4%) | 60/110 (54.5%) | 0.085 |
| Mid-upper arm circumference, cm | 243 | 24 (22, 26) | 24 (22, 26) | 24 (22, 26) | 0.3 |
| Illness duration prior to enrollment, days | 243 | 7 (4, 11) | 5 (4, 8) | 8 (5, 13) | <0.001 |
| Lactate, mmol/L | 243 | 2.4 (1.8, 3.1) | 2.2 (1.8, 2.7) | 2.8 (1.9, 3.6) | <0.001 |
| Lactate ≥ 4 mmol/L | 243 | 24/243 (9.9%) | 7/133 (5.3%) | 17/110 (15.5%) | 0.008 |
| Modified Early Warning Score | 243 | 4 (3, 5) | 3 (3, 4) | 4 (3, 6) | <0.001 |
| Universal Vital Assessment score | 243 | 3 (1, 6) | 3 (1, 5) | 4 (2, 6) | <0.001 |
| Living with HIV | 240 | 119/240 (49.6%) | 61/131 (46.6%) | 58/109 (53.2%) | 0.3 |
| Malaria RDT positive | 243 | 70/243 (28.8%) | 45/133 (33.8%) | 25/110 (22.7%) | 0.057 |
| Microbiological TB positive | 243 | 51/243 (21.0%) | 25/133 (18.8%) | 26/110 (23.6%) | 0.4 |
| Death by 60 days | 241 | 74/241 (30.7%) | 29/132 (22.0%) | 45/109 (41.3%) | 0.001 |
| *^1^*Median (Q1, Q3); n/N (%) | | | | | |
| *^2^*Wilcoxon rank sum test; Pearson's Chi-squared test | | | | | |

| **Characteristic** | **N** | **Overall**  **N = 112*^1^*** | **USE-1**  **N = 77*^1^*** | **USE-2**  **N = 35*^1^*** | **p-value*^2^*** |
| --- | --- | --- | --- | --- | --- |
| Age, years | 112 | 33 (27, 43) | 35 (28, 43) | 30 (25, 43) | 0.2 |
| Female sex | 112 | 70/112 (62.5%) | 49/77 (63.6%) | 21/35 (60.0%) | 0.7 |
| Illness duration prior to enrollment, days | 111 | 5 (4, 8), 111 | 5 (4, 8), 77 | 6 (4, 8), 34 | 0.4 |
| Modified Early Warning Score | 112 | 3 (2, 5) | 3 (2, 4) | 4 (3, 6) | 0.003 |
| Universal Vital Assessment score | 112 | 3 (2, 4) | 2 (2, 4) | 4 (2, 5) | 0.063 |
| Living with HIV | 111 | 63/111 (56.8%) | 39/76 (51.3%) | 24/35 (68.6%) | 0.088 |
| Malaria RDT positive | 108 | 21/108 (19.4%) | 17/75 (22.7%) | 4/33 (12.1%) | 0.2 |
| Microbiological TB positive | 112 | 19/112 (17.0%) | 9/77 (11.7%) | 10/35 (28.6%) | 0.027 |
| Death by 30 days | 104 | 29/104 (27.9%) | 16/72 (22.2%) | 13/32 (40.6%) | 0.053 |
| *^1^*n/N (%); Median (Q1, Q3) | | | | | |
| *^2^*Pearson's Chi-squared test; Wilcoxon rank sum test | | | | | |

**Table S2: Patient characteristics stratified by Uganda Sepsis Endotypes (USE) in RESERVE-U-1-EBB**

**Table S3: Patient characteristics stratified by Uganda Sepsis Signatures (USS) in RESERVE-U-2-TOR**

| **Characteristic** | **N** | **Overall**  **N = 253*^1^*** | **USS-1**  **N = 190*^1^*** | **USS-2**  **N = 63*^1^*** | **p-value*^2^*** |
| --- | --- | --- | --- | --- | --- |
| Age, years | 253 | 47 (34, 61) | 48 (33, 60) | 47 (36, 65) | 0.6 |
| Female sex | 253 | 152/253 (60.1%) | 121/190 (63.7%) | 31/63 (49.2%) | 0.042 |
| Mid-upper arm circumference, cm | 253 | 24 (22, 26) | 24 (22, 26) | 24 (22, 26) | 0.7 |
| Illness duration prior to enrollment, days | 253 | 6 (4, 11) | 6 (4, 9) | 9 (6, 13) | <0.001 |
| Lactate, mmol/L | 253 | 2.4 (1.9, 3.1) | 2.4 (1.9, 3.0) | 2.6 (1.8, 3.7) | 0.2 |
| Lactate ≥ 4 mmol/L | 253 | 26/253 (10.3%) | 15/190 (7.9%) | 11/63 (17.5%) | 0.030 |
| Modified Early Warning Score | 253 | 4 (3, 5) | 4 (3, 5) | 4 (3, 6) | 0.011 |
| Universal Vital Assessment score | 253 | 3 (2, 6) | 3 (1, 5) | 4 (2, 6) | 0.047 |
| Living with HIV | 250 | 123/250 (49.2%) | 90/188 (47.9%) | 33/62 (53.2%) | 0.5 |
| Malaria RDT positive | 253 | 72/253 (28.5%) | 56/190 (29.5%) | 16/63 (25.4%) | 0.5 |
| Microbiological TB positive | 253 | 52/253 (20.6%) | 35/190 (18.4%) | 17/63 (27.0%) | 0.14 |
| Death by 60 days | 251 | 77/251 (30.7%) | 48/189 (25.4%) | 29/62 (46.8%) | 0.002 |
| *^1^*Median (Q1, Q3); n/N (%) | | | | | |
| *^2^*Wilcoxon rank sum test; Pearson's Chi-squared test | | | | | |

**Table S4: Patient characteristics stratified by Uganda Sepsis Signatures (USS) in RESERVE-U-1-EBB**

| **Characteristic** | **N** | **Overall**  **N = 242*^1^*** | **USS-1**  **N = 156*^1^*** | **USS-2**  **N = 86*^1^*** | **p-value*^2^*** |
| --- | --- | --- | --- | --- | --- |
| Age, years | 242 | 32 (26, 43) | 31 (26, 40) | 36 (28, 46) | 0.009 |
| Female sex | 242 | 143/242 (59.1%) | 92/156 (59.0%) | 51/86 (59.3%) | >0.9 |
| Illness duration prior to enrollment, days | 241 | 5 (4, 8), 241 | 5 (3, 8), 156 | 6 (4, 8), 85 | 0.12 |
| Modified Early Warning Score | 242 | 3 (2, 5) | 3 (2, 4) | 4 (3, 6) | <0.001 |
| Universal Vital Assessment score | 242 | 3 (2, 4) | 2 (1, 4) | 4 (2, 6) | <0.001 |
| Living with HIV | 240 | 136/240 (56.7%) | 70/154 (45.5%) | 66/86 (76.7%) | <0.001 |
| Malaria RDT positive | 238 | 48/238 (20.2%) | 35/155 (22.6%) | 13/83 (15.7%) | 0.2 |
| Microbiological TB positive | 242 | 47/242 (19.4%) | 22/156 (14.1%) | 25/86 (29.1%) | 0.005 |
| Death by 30 days | 220 | 59/220 (26.8%) | 27/144 (18.8%) | 32/76 (42.1%) | <0.001 |
| *^1^*n/N (%); Median (Q1, Q3) | | | | | |
| *^2^*Pearson's Chi-squared test; Wilcoxon rank sum test | | | | | |

**Table S5: Patient characteristics stratified by Consensus Transcriptomic Subtypes (CTS) in RESERVE-U-2-TOR**

| **Characteristic** | **N** | **Overall**  **N = 243*^1^*** | **CTS1**  **N = 82*^1^*** | **CTS2**  **N = 58*^1^*** | **CTS3**  **N = 103*^1^*** | **p-value*^2^*** |
| --- | --- | --- | --- | --- | --- | --- |
| Age, years | 243 | 48 (35, 62) | 50 (42, 67) | 43 (30, 60) | 48 (30, 60) | 0.041 |
| Female sex | 243 | 147/243 (60.5%) | 43/82 (52.4%) | 37/58 (63.8%) | 67/103 (65.0%) | 0.2 |
| Mid-upper arm circumference, cm | 243 | 24 (22, 26) | 24 (22, 26) | 24 (22, 26) | 24 (22, 26) | 0.3 |
| Illness duration prior to enrollment, days | 243 | 7 (4, 11) | 7 (4, 12) | 8 (5, 13) | 5 (4, 8) | 0.001 |
| Whole-blood lactate, mmol/L | 243 | 2.4 (1.8, 3.1) | 2.5 (1.9, 3.3) | 2.6 (1.9, 3.4) | 2.2 (1.6, 2.8) | 0.001 |
| Whole-blood lactate ≥ 4 mmol/L | 243 | 24/243 (9.9%) | 9/82 (11.0%) | 10/58 (17.2%) | 5/103 (4.9%) | 0.038 |
| Modified Early Warning Score | 243 | 4 (3, 5) | 4 (3, 6) | 4 (3, 5) | 3 (3, 5) | 0.019 |
| Universal Vital Assessment score | 243 | 3 (1, 6) | 4 (2, 6) | 3 (2, 5) | 3 (1, 5) | 0.2 |
| Living with HIV | 240 | 119/240 (49.6%) | 40/81 (49.4%) | 29/57 (50.9%) | 50/102 (49.0%) | >0.9 |
| Malaria RDT positive | 243 | 70/243 (28.8%) | 22/82 (26.8%) | 15/58 (25.9%) | 33/103 (32.0%) | 0.6 |
| Microbiological TB positive | 243 | 51/243 (21.0%) | 17/82 (20.7%) | 13/58 (22.4%) | 21/103 (20.4%) | >0.9 |
| Death by 60 days | 241 | 74/241 (30.7%) | 31/81 (38.3%) | 22/57 (38.6%) | 21/103 (20.4%) | 0.011 |
| *^1^*Median (Q1, Q3); n/N (%) | | | | | | |
| *^2^*Kruskal-Wallis rank sum test; Pearson's Chi-squared test | | | | | | |

**Table S6: Patient characteristics stratified by Consensus Transcriptomic Subtypes (CTS) in RESERVE-U-1-EBB**

| **Characteristic** | **N** | **Overall**  **N = 112*^1^*** | **CTS1**  **N = 38*^1^*** | **CTS2**  **N = 30*^1^*** | **CTS3**  **N = 44*^1^*** | **p-value*^2^*** |
| --- | --- | --- | --- | --- | --- | --- |
| Age, years | 112 | 33 (27, 43) | 35 (28, 46) | 31 (25, 40) | 32 (27, 43) | 0.4 |
| Female sex | 112 | 70/112 (62.5%) | 24/38 (63.2%) | 19/30 (63.3%) | 27/44 (61.4%) | >0.9 |
| Illness duration prior to enrollment, days | 111 | 5 (4, 8), 111 | 6 (4, 8), 37 | 5 (4, 8), 30 | 5 (4, 8), 44 | 0.8 |
| Modified Early Warning Score | 112 | 3 (2, 5) | 4 (3, 6) | 3 (2, 5) | 3 (2, 4) | 0.001 |
| Universal Vital Assessment score | 112 | 3 (2, 4) | 3 (2, 5) | 3 (2, 4) | 2 (1, 4) | 0.022 |
| Living with HIV | 111 | 63/111 (56.8%) | 29/37 (78.4%) | 12/30 (40.0%) | 22/44 (50.0%) | 0.004 |
| Malaria RDT positive | 108 | 21/108 (19.4%) | 6/35 (17.1%) | 5/29 (17.2%) | 10/44 (22.7%) | 0.8 |
| Microbiological TB positive | 112 | 19/112 (17.0%) | 11/38 (28.9%) | 4/30 (13.3%) | 4/44 (9.1%) | 0.048 |
| Death by 30 days | 104 | 29/104 (27.9%) | 15/35 (42.9%) | 6/28 (21.4%) | 8/41 (19.5%) | 0.052 |
| *^1^*n/N (%); Median (Q1, Q3) | | | | | | |
| *^2^*Pearson's Chi-squared test; Kruskal-Wallis rank sum test | | | | | | |

**Table S7: Patient characteristics stratified by Hi-DEF Myeloid Dysregulation Score Tertiles in RESERVE-U-2-TOR**

| **Characteristic** | **N** | **Overall**  **N = 243*^1^*** | **Low Myeloid**  **N = 81*^1^*** | **Mid Myeloid**  **N = 81*^1^*** | **High Myeloid**  **N = 81*^1^*** | **p-value*^2^*** |
| --- | --- | --- | --- | --- | --- | --- |
| Age, years | 243 | 48 (35, 62) | 46 (32, 54) | 45 (29, 63) | 55 (43, 66) | 0.003 |
| Female sex | 243 | 147/243 (60.5%) | 49/81 (60.5%) | 52/81 (64.2%) | 46/81 (56.8%) | 0.6 |
| Mid-upper arm circumference, cm | 243 | 24 (22, 26) | 24 (22, 26) | 24 (22, 26) | 24 (22, 26) | 0.5 |
| Illness duration prior to enrollment, days | 243 | 7 (4, 11) | 6 (3, 8) | 6 (4, 9) | 8 (5, 13) | <0.001 |
| Whole-blood lactate, mmol/L | 243 | 2.4 (1.8, 3.1) | 2.1 (1.6, 2.7) | 2.5 (2.1, 3.3) | 2.5 (1.9, 3.1) | 0.003 |
| Whole-blood lactate ≥ 4 mmol/L | 243 | 24/243 (9.9%) | 4/81 (4.9%) | 9/81 (11.1%) | 11/81 (13.6%) | 0.2 |
| Modified Early Warning Score | 243 | 4 (3, 5) | 4 (3, 5) | 4 (3, 5) | 4 (3, 5) | 0.3 |
| Universal Vital Assessment score | 243 | 3 (1, 6) | 2 (1, 5) | 3 (1, 5) | 3 (2, 6) | 0.077 |
| Living with HIV | 240 | 119/240 (49.6%) | 42/79 (53.2%) | 34/80 (42.5%) | 43/81 (53.1%) | 0.3 |
| Malaria RDT positive | 243 | 70/243 (28.8%) | 23/81 (28.4%) | 28/81 (34.6%) | 19/81 (23.5%) | 0.3 |
| Microbiological TB positive | 243 | 51/243 (21.0%) | 15/81 (18.5%) | 17/81 (21.0%) | 19/81 (23.5%) | 0.7 |
| Death by 60 days | 241 | 74/241 (30.7%) | 19/81 (23.5%) | 24/81 (29.6%) | 31/79 (39.2%) | 0.093 |
| *^1^*Median (Q1, Q3); n/N (%) | | | | | | |
| *^2^* Pearson's Chi-squared test; Kruskal-Wallis rank sum test. | | | | | | |

**Table S8: Patient characteristics stratified by Hi-DEF Myeloid Dysregulation Score Tertiles in RESERVE-U-1-EBB**

| **Characteristic** | **N** | **Overall**  **N = 112*^1^*** | **Low Myeloid**  **N = 38*^1^*** | **Mid Myeloid**  **N = 37*^1^*** | **High Myeloid**  **N = 37*^1^*** | **p-value*^2^*** |
| --- | --- | --- | --- | --- | --- | --- |
| Age, years | 112 | 33 (27, 43) | 37 (28, 46) | 30 (27, 43) | 31 (25, 40) | 0.15 |
| Female sex | 112 | 70/112 (62.5%) | 24/38 (63.2%) | 21/37 (56.8%) | 25/37 (67.6%) | 0.6 |
| Illness duration prior to enrollment, days | 111 | 5 (4, 8), 111 | 6 (4, 8), 38 | 5 (3, 7), 37 | 6 (4, 8), 36 | 0.2 |
| Modified Early Warning Score | 112 | 3 (2, 5) | 3 (2, 5) | 3 (2, 5) | 3 (2, 5) | 0.7 |
| Universal Vital Assessment score | 112 | 3 (2, 4) | 3 (2, 4) | 2 (2, 4) | 3 (2, 4) | 0.6 |
| Living with HIV | 111 | 63/111 (56.8%) | 23/38 (60.5%) | 20/37 (54.1%) | 20/36 (55.6%) | 0.8 |
| Malaria RDT positive | 108 | 21/108 (19.4%) | 6/37 (16.2%) | 10/36 (27.8%) | 5/35 (14.3%) | 0.3 |
| Microbiological TB positive | 112 | 19/112 (17.0%) | 8/38 (21.1%) | 5/37 (13.5%) | 6/37 (16.2%) | 0.7 |
| Death by 30 days | 104 | 29/104 (27.9%) | 9/35 (25.7%) | 11/34 (32.4%) | 9/35 (25.7%) | 0.8 |
| *^1^*n/N (%); Median (Q1, Q3) | | | | | | |
| *^2^*Pearson's Chi-squared test; Kruskal-Wallis rank sum test. | | | | | | |

**Table S9: Patient characteristics stratified by Hi-DEF Lymphoid Dysregulation Score Tertiles in RESERVE-U-2-TOR**

| **Characteristic** | **N** | **Overall**  **N = 243*^1^*** | **Low Lymphoid**  **N = 81*^1^*** | **Mid**  **Lymphoid**  **N = 81*^1^*** | **High Lymphoid**  **N = 81*^1^*** | **p-value*^2^*** |
| --- | --- | --- | --- | --- | --- | --- |
| Age, years | 243 | 48 (35, 62) | 47 (28, 58) | 50 (34, 64) | 47 (39, 69) | 0.2 |
| Female sex | 243 | 147/243 (60.5%) | 60/81 (74.1%) | 45/81 (55.6%) | 42/81 (51.9%) | 0.008 |
| Mid-upper arm circumference, cm | 243 | 24 (22, 26) | 24 (23, 26) | 23 (22, 26) | 24 (22, 26) | 0.13 |
| Illness duration prior to enrollment, days | 243 | 7 (4, 11) | 5 (4, 8) | 7 (4, 13) | 7 (5, 13) | 0.007 |
| Whole-blood lactate, mmol/L | 243 | 2.4 (1.8, 3.1) | 2.3 (1.8, 2.8) | 2.4 (1.9, 3.0) | 2.7 (1.8, 3.6) | 0.036 |
| Whole-blood lactate ≥ 4 mmol/L | 243 | 24/243 (9.9%) | 5/81 (6.2%) | 7/81 (8.6%) | 12/81 (14.8%) | 0.2 |
| Modified Early Warning Score | 243 | 4 (3, 5) | 3 (3, 5) | 4 (3, 4) | 5 (3, 6) | <0.001 |
| Universal Vital Assessment score | 243 | 3 (1, 6) | 3 (1, 5) | 3 (2, 6) | 4 (2, 6) | 0.053 |
| Living with HIV | 240 | 119/240 (49.6%) | 33/79 (41.8%) | 48/81 (59.3%) | 38/80 (47.5%) | 0.078 |
| Malaria RDT positive | 243 | 70/243 (28.8%) | 24/81 (29.6%) | 31/81 (38.3%) | 15/81 (18.5%) | 0.021 |
| Microbiological TB positive | 243 | 51/243 (21.0%) | 11/81 (13.6%) | 23/81 (28.4%) | 17/81 (21.0%) | 0.069 |
| Death by 60 days | 241 | 74/241 (30.7%) | 15/80 (18.8%) | 27/81 (33.3%) | 32/80 (40.0%) | 0.012 |
| *^1^*Median (Q1, Q3); n/N (%) | | | | | | |
| *^2^*Pearson's Chi-squared test; Kruskal-Wallis rank sum test. | | | | | | |

**Table S10: Patient characteristics stratified by Hi-DEF Lymphoid Dysregulation Score Tertiles in RESERVE-U-1-EBB**

| **Characteristic** | **N** | **Overall**  **N = 112*^1^*** | **Low Lymphoid**  **N = 38*^1^*** | **Mid Lymphoid**  **N = 37*^1^*** | **High Lymphoid**  **N = 37*^1^*** | **p-value*^2^*** |
| --- | --- | --- | --- | --- | --- | --- |
| Age, years | 112 | 33 (27, 43) | 31 (26, 45) | 33 (27, 43) | 33 (29, 43) | 0.8 |
| Female sex | 112 | 70/112 (62.5%) | 25/38 (65.8%) | 22/37 (59.5%) | 23/37 (62.2%) | 0.9 |
| Illness duration prior to enrollment, days | 111 | 5 (4, 8), 111 | 5 (3, 6), 38 | 6 (4, 8), 37 | 6 (4, 8), 36 | 0.063 |
| Modified Early Warning Score | 112 | 3 (2, 5) | 3 (2, 3) | 4 (3, 5) | 4 (3, 6) | <0.001 |
| Universal Vital Assessment score | 112 | 3 (2, 4) | 2 (1, 4) | 3 (2, 4) | 4 (2, 5) | 0.006 |
| Living with HIV | 111 | 63/111 (56.8%) | 10/38 (26.3%) | 26/37 (70.3%) | 27/36 (75.0%) | <0.001 |
| Malaria RDT positive | 108 | 21/108 (19.4%) | 9/37 (24.3%) | 9/37 (24.3%) | 3/34 (8.8%) | 0.2 |
| Microbiological TB positive | 112 | 19/112 (17.0%) | 2/38 (5.3%) | 5/37 (13.5%) | 12/37 (32.4%) | 0.006 |
| Death by 30 days | 104 | 29/104 (27.9%) | 4/35 (11.4%) | 9/34 (26.5%) | 16/35 (45.7%) | 0.006 |
| *^1^*n/N (%); Median (Q1, Q3) | | | | | | |
| *^2^*Pearson's Chi-squared test; Kruskal-Wallis rank sum test. | | | | | | |

**Table S11: Performance characteristics of clinical and clinico-microbiological models for prediction of molecular sepsis subtypes and scores in RESERVE-U-2-TOR**

| **Models and Outcomes^a^** | **AUROC**  **(95% CI)^b^** | **Integrated Calibration Index (E_avg_)^c^** | **Pearson *r***  **(95% CI)^b^** | **Mean Absolute Error** |
| --- | --- | --- | --- | --- |
| **Uganda Sepsis Endotypes (Transcriptomic)** | | | | |
| Clinical^d^ | 0.753  (0.691-0.811) | 0.014 | -- | -- |
| Clinical + Microbiological^e^ | 0.756  (0.694-0.814) | 0.016 | -- | -- |
| **Uganda Sepsis Signatures (Proteomic)** | | | | |
| Clinical^f^ | 0.728  (0.655-0.798) | 0.010 | -- | -- |
| Clinical + Microbiological^g^ | 0.739  (0.665-0.809) | 0.013 | -- | -- |
| **Consensus Transcriptomic Subtypes (Transcriptomic)** | | | | |
| Clinical^d^ | 0.679  (0.628-0.730) | 0.019 | -- | -- |
| Clinical + Microbiological^e^ | 0.683  (0.631-0.733) | 0.018 | -- | -- |
| **Hi-DEF Myeloid Dysregulation Score (Transcriptomic)** | | | | |
| Clinical^d^ | -- | -- | 0.276  (0.128-0.413) | 1.635 |
| Clinical + Microbiological^e^ | -- | -- | 0.291  (0.145-0.425) | 1.661 |
| **Hi-DEF Lymphoid Dysregulation Score (Transcriptomic)** | | | | |
| Clinical^d^ | -- | -- | 0.518  (0.410-0.612) | 0.228 |
| Clinical + Microbiological^e^ | -- | -- | 0.520  (0.409-0.613) | 0.228 |

Abbreviations: AUROC: area under the receiver operating characteristic curve; CI: Confidence interval.

Legend: ^a^Clinical model includes age (continuous, years), sex (binary), duration of illness prior to presentation (continuous, days), temperature (continuous, **°**C**)**, heart rate (continuous, beats/min), respiratory rate (continuous, breaths/min), systolic blood pressure (continuous, mmHg), oxygen saturation (continuous, %), and mental status assessed by AVPU (categorical, Alert, Responsive to Voice, Responsive to Pain, Unresponsive); Clinical + Microbiological model includes clinical model variables plus results of rapid diagnostics for HIV (binary), malaria (binary), and tuberculosis (binary); ^b^95% CIs generated using 10,000 bootstrapped replicates; ^c^Integrated Calibration Index (E_avg_) indicates average absolute difference between predicted probabilities and LOESS-smoothed observed probabilities; ^d^N=243, ^e^N=240, ^f^N=253, ^g^N=250.

**Table S12: Performance characteristics of clinical and clinico-microbiological models for prediction of molecular sepsis subtypes and scores in RESERVE-U-1-EBB**

| **Models and Outcomes^a^** | **AUROC**  **(95% CI)^b^** | **Integrated Calibration Index (E_avg_)^c^** | **Pearson *r***  **(95% CI)^b^** | **Mean Absolute Error** |
| --- | --- | --- | --- | --- |
| **Uganda Sepsis Endotypes (Transcriptomic)** | | | | |
| Clinical^d^ | 0.634  (0.497-0.792) | 0.053 | -- | -- |
| Clinical + Microbiological^e^ | 0.688  (0.561-0.804) | 0.039 | -- | -- |
| **Uganda Sepsis Signatures (Proteomic)** | | | | |
| Clinical^f^ | 0.720  (0.646-0.788) | 0.026 | -- | -- |
| Clinical + Microbiological^g^ | 0.751  (0.682-0.816) | 0.017 | -- | -- |
| **Consensus Transcriptomic Subtypes (Transcriptomic)** | | | | |
| Clinical^d^ | 0.689  (0.613-0.764) | 0.030 | -- | -- |
| Clinical + Microbiological^e^ | 0.743  (0.671-0.810) | 0.030 | -- | -- |
| **Hi-DEF Myeloid Dysregulation Score (Transcriptomic)** | | | | |
| Clinical^d^ | -- | -- | 0.275  (0.082-0.452) | 1.527 |
| Clinical + Microbiological^e^ | -- | -- | 0.259  (0.066-0.431) | 1.523 |
| **Hi-DEF Lymphoid Dysregulation Score (Transcriptomic)** | | | | |
| Clinical^d^ | -- | -- | 0.397  (0.196-0.574) | 0.421 |
| Clinical + Microbiological^e^ | -- | -- | 0.487  (0.298-0.654) | 0.388 |

Abbreviations: AUROC: area under the receiver operating characteristic curve; CI: Confidence interval.

Legend: ^a^Clinical model includes age (continuous, years), sex (binary), duration of illness prior to presentation (continuous, days), temperature (continuous, **°**C**)**, heart rate (continuous, beats/min), respiratory rate (continuous, breaths/min), systolic blood pressure (continuous, mmHg), oxygen saturation (continuous, %), and mental status assessed by AVPU (categorical, Alert, Responsive to Voice, Responsive to Pain, Unresponsive); Clinical + Microbiological model includes clinical model variables plus results of rapid diagnostics for HIV (binary), malaria (binary), and tuberculosis (binary); ^b^95% CIs generated using 10,000 bootstrapped replicates; ^c^Integrated Calibration Index (E_avg_) indicates average absolute difference between predicted probabilities and LOESS-smoothed observed probabilities; ^d^N=111, ^e^N=106, ^f^N=241, ^g^N=235.
